# Targeted malaria elimination interventions reduce *Plasmodium falciparum* infections up to 3 kilometers away

**DOI:** 10.1101/2023.09.19.23295806

**Authors:** Jade Benjamin-Chung, Haodong Li, Anna Nguyen, Gabriella Barratt Heitmann, Adam Bennett, Henry Ntuku, Lisa M. Prach, Munyaradzi Tambo, Lindsey Wu, Chris Drakeley, Roly Gosling, Davis Mumbengegwi, Immo Kleinschmidt, Jennifer L. Smith, Alan Hubbard, Mark van der Laan, Michelle S. Hsiang

## Abstract

Malaria elimination interventions in low-transmission settings aim to extinguish hot spots and prevent transmission to nearby areas. In malaria elimination settings, the World Health Organization recommends reactive, focal interventions targeted to the area near malaria cases shortly after they are detected. A key question is whether these interventions reduce transmission to nearby uninfected or asymptomatic individuals who did not receive interventions. Here, we measured direct effects (among intervention recipients) and spillover effects (among non-recipients) of reactive, focal interventions delivered within 500m of confirmed malaria index cases in a cluster-randomized trial in Namibia. The trial delivered malaria chemoprevention (artemether lumefantrine) and vector control (indoor residual spraying with Actellic) separately and in combination using a factorial design. We compared incidence, infection prevalence, and seroprevalence between study arms among intervention recipients (direct effects) and non-recipients (spillover effects) up to 3 km away from index cases. We calculated incremental cost-effectiveness ratios accounting for spillover effects. The combined chemoprevention and vector control intervention produced direct effects and spillover effects. In the primary analysis among non-recipients within 1 km from index cases, the combined intervention reduced malaria incidence by 43% (95% CI 20%, 59%). In secondary analyses among non-recipients 500m-3 km from interventions, the combined intervention reduced infection by 79% (6%, 95%) and seroprevalence 34% (20%, 45%). Accounting for spillover effects increased the cost-effectiveness of the combined intervention by 37%. Our findings provide the first evidence that targeting hot spots with combined chemoprevention and vector control interventions can indirectly benefit non-recipients up to 3 km away.

**Significance Statement:** In settings where malaria transmission is declining and approaching elimination, new malaria cases are clustered in space and time. Prior studies have found that targeting prophylactic antimalarial drugs and vector control in the area around newly detected malaria cases reduced community-wide malaria. Here, we found that when antimalarials and vector control were delivered as a combined strategy in the area near recent cases, malaria incidence was reduced up to 3 kilometers away among individuals who did not receive interventions. Accounting for these benefits to non-recipients increased cost-effectiveness of the intervention. Overall, our findings suggest that combined, targeted malaria interventions can reduce local transmission and support their use for malaria elimination.

## Introduction

In the past decade, there has been renewed attention towards global malaria eradication, and many countries have set targets for the elimination of local malaria transmission (1). In Southern Africa, eight countries hope to achieve malaria elimination by 2030 as part of the Elimination Eight Initiative (E8). Yet, global progress has plateaued: annual global malaria cases have increased since 2015, and in 2021, there were 59 cases per 1,000 population at risk, an increase from 57 per 1,000 in 2019 (2).

The ideal malaria elimination intervention would not only prevent disease among recipients but would also prevent onward transmission to nearby non-recipients through spillover effects (i.e., “herd effects”, “indirect effects”) (4, 5), like some vaccines do (4, 6–12). Prior studies have reported spillover effects for mass drug administration for trachoma (13, 14), school-based deworming (15), insecticide treated bed nets (16–18), and chemoprevention and vector control for malaria (19, 20).

When an intervention reduces disease among intervention non-recipients, accounting for spillover effects can substantially increase cost-effectiveness (21, 22). Identifying cost-effective interventions is crucial to the elimination and eradication enterprise because elimination efforts are projected to cost significantly more than existing malaria control programs in the medium term (3). Even after elimination, countries must continue to conduct intensive surveillance and outbreak response for imported cases to prevent re-establishment.

In settings approaching malaria elimination, the World Health Organization recommends interventions that are “reactive” – delivered soon after a confirmed malaria case is detected – and “focal” – delivered to higher risk individuals who reside near the case (23). A recent cluster-randomized trial in Namibia found that reactive, focal chemoprevention and vector control substantially reduced malaria incidence (24). Spillover effects of these interventions are plausible: chemoprevention may reduce gametocyte biomass in recipients (25), and vector control can reduce the mosquito population near malaria cases. To shed light on whether focal interventions reduce transmission to nearby uninfected or asymptomatic individuals who did not receive interventions, we separately estimated direct effects among intervention recipients and spillover effects among nearby non-recipients. Our approach can be used to estimate spillover effects of other interventions, such as malaria vaccines.

## Results

### Interventions

We analyzed data from a previously reported cluster-randomized trial of focal malaria interventions conducted in Zambezi region of Namibia in 2017 (NCT02610400) (24) (Figure 1). The region has low *Plasmodium falciparum* malaria transmission (26). Using a two-by-two factorial design, the trial randomized 56 clusters to four arms: 1) reactive case detection (RACD), 2) reactive focal mass drug administration (rfMDA) only, 3) reactive vector control (RAVC) + RACD, 4) RAVC + rfMDA. rfMDA included presumptive treatment with artemether-lumefrantrine to individuals in target areas (Table S1). RACD included testing with rapid diagnostic tests and treatment with artemether-lumefrantrine and single-dose primaquine for those who tested positive. RAVC included indoor residual spraying (IRS) with pirimiphosmethyl. The trial delivered interventions in “target areas” within approximately 500 m of confirmed malaria cases detected through passive surveillance.

**Figure 1.**
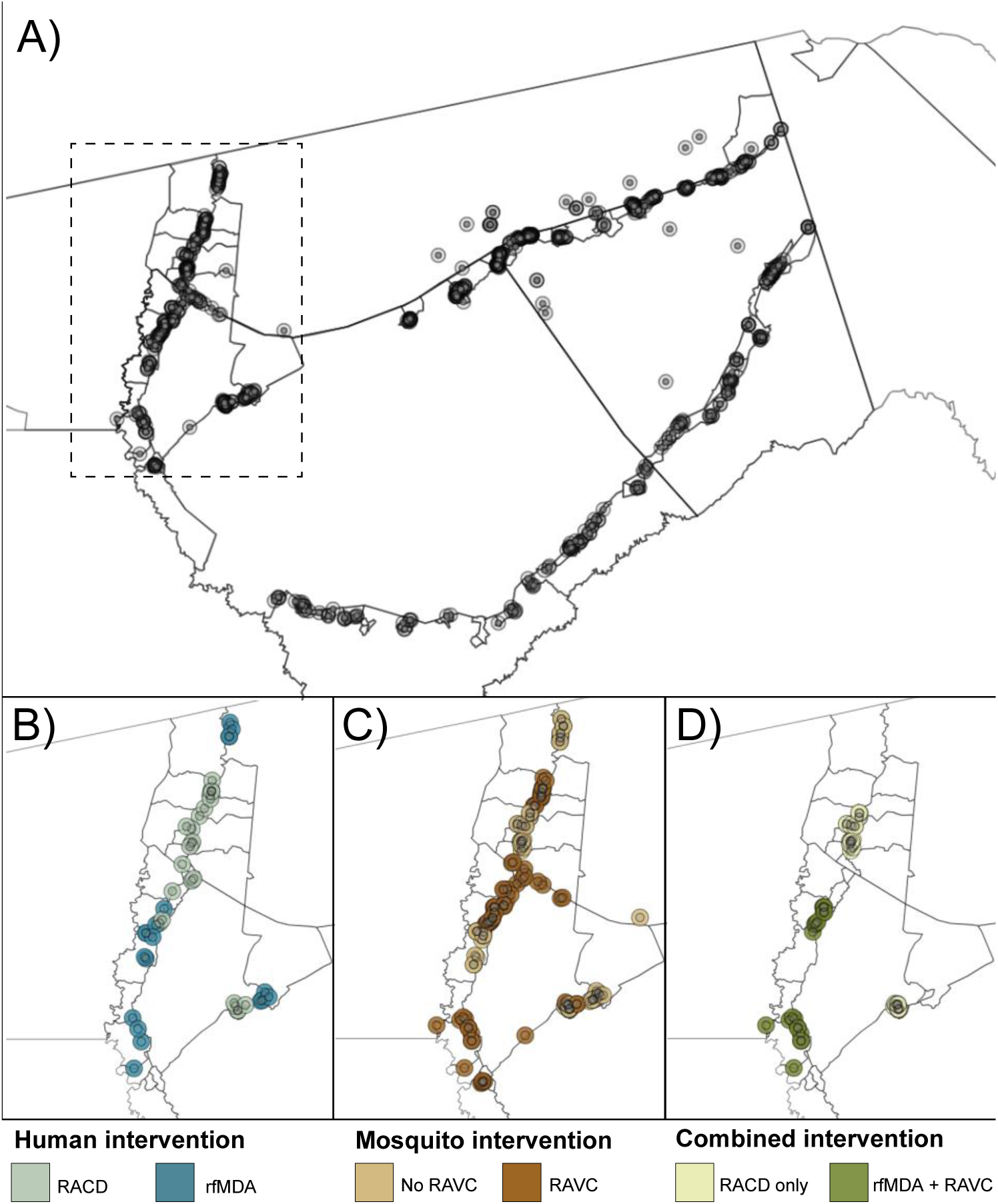
Map of target areas and spillover zones in the study site. **a)** All index cases during the study period. The centroid of each circle is the residence location of a treated index case. Inner circles indicate 500m target areas where interventions were delivered. Outer circles indicate the 1km radius around each index case in which spillover effects were estimated in primary analyses. The dashed line indicates insets in panels B-D showing index cases during the follow-up periods with the largest number of treated index cases. For each comparison of study arms, panels b-d depict examples of analytic cohorts from a single follow-up period (i.e., subsample of person-time) in a subset of the study area. **b**) Inset of study area with index cases in the RACD and rfMDA arms (5-week period: April 25, 2017 – May 30, 2017). **C**) Inset of study area with index cases in the no RAVC and RAVC arms (6-month period: January 1, 2017 - June 30, 2017). **d**) Inset of study area with index cases in the RAVC and rfMDA+RAVC arms (6-month period: January 1, 2017 - June 30, 2017). RACD: reactive case detection rfMDA: reactive focal mass drug administration RAVC: reactive focal vector control

### Effects on malaria incidence

Primary analyses estimated effects on malaria incidence. We estimated three types of effects on the cumulative incidence of locally acquired, confirmed *Pf* malaria: 1) direct effects among intervention recipients in target areas within 500 m of confirmed malaria index cases, 2) spillover effects among non-recipients within 1 km of index cases, and 3) total effects among all individuals within 1 km of index cases (Figure 2a). We created analytic cohorts including individuals residing within 1 km of each index case to capture the area and time period in which we expected each intervention to reduce infections (direct effect) and secondary transmission to nearby individuals (spillover effect). (Figure 1; Figure S1 see details in Materials and Methods). We measured effects of the chemoprevention intervention comparing arms with rfMDA vs. RACD, the vector control intervention by comparing arms with RAVC vs. no RAVC, and the combined intervention by comparing the rfMDA + RAVC vs. RACD only arms.

**Figure 2.**
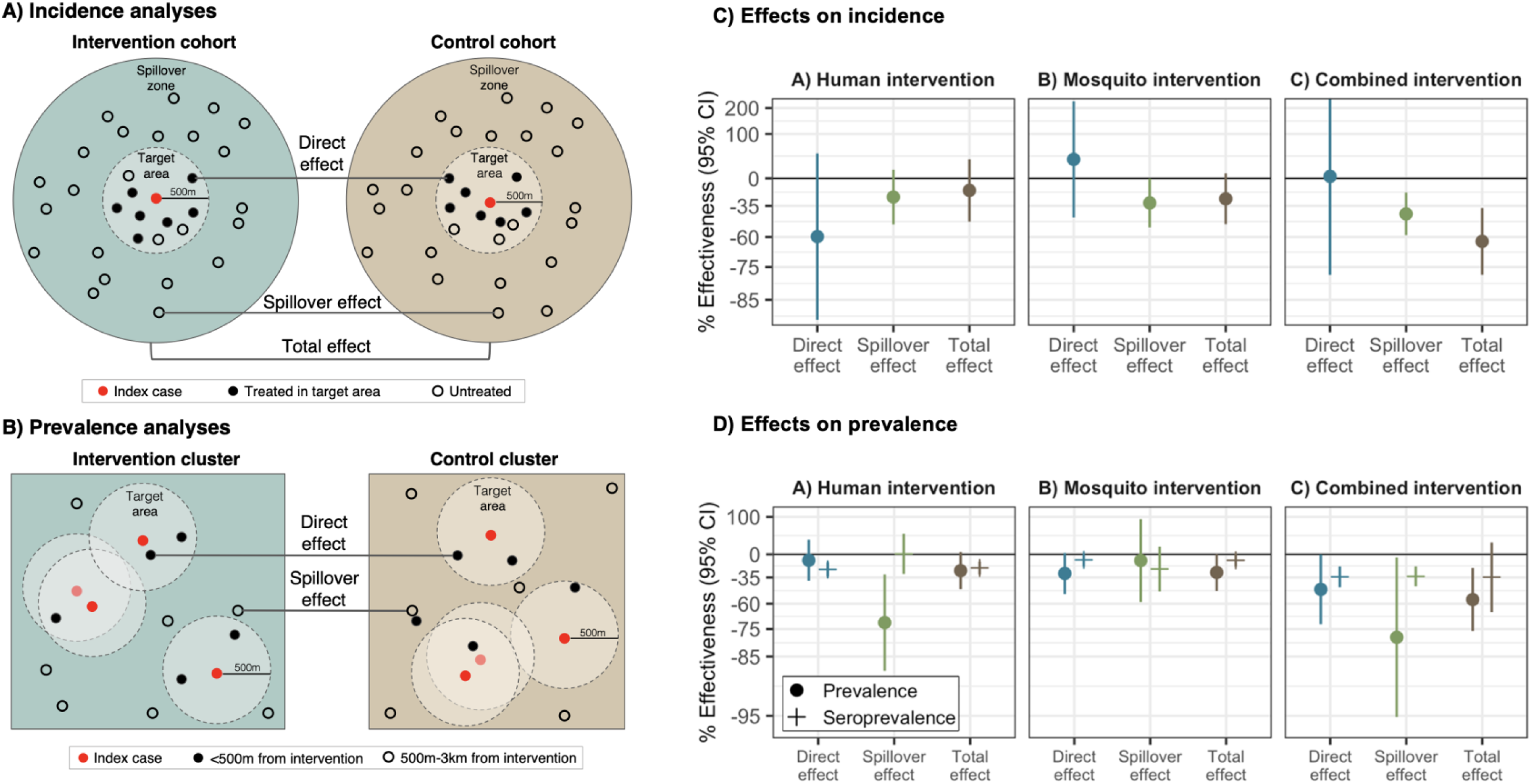
Effects of reactive, focal malaria interventions. **a**) Definition of effects in incidence analyses. **b**) Definition of effects in prevalence analyses. **c**) Effects on incidence estimated with hierarchical TMLE. All incidence outcome models were fit with cohort-level data except for models of spillover effects of rfMDA vs. RACD and rfMDA + RAVC vs. RACD only. Models were adjusted for covariates that were screened separately for each model using a likelihood ratio test. Confidence intervals shown here do not account for potential outcome correlation. For rfMDA and RACD arms, the incidence analysis includes the period from 0-35 days following index case detection for direct effects and 21-56 days for spillover effects. For rfMDA+RAVC and RAVC only arms, the analysis includes the period from 0-6 months following index case detection for direct effects and 17 days to 6 months for spillover effects. Total effects analyses include the person-time for the direct effects and spillover effects analyses. For incidence analyses, direct effects include treated in target zone, spillover effects include intervention non-recipients up to 1km from an index case, and total effects include all individuals (intervention recipients and non-recipients) up to 1km from index case. **d**) Effects on prevalence estimated with TMLE using individual-level data; standard errors were adjusted for clustering at the enumeration area level. Models were unadjusted because there were fewer than 10 malaria cases per variable. Direct effects include individuals who resided within 500m of any intervention recipients, spillover effects include individuals with no intervention recipients < 500m and any intervention recipients 500m-3km, and total effects include individuals with any intervention recipients <3km during the study. In **c**) and **d**), % effectiveness was calculated as the ratio of incidence or prevalence between study arms minus 1 x 100%. The upper bound of the 95% CI for the combined intervention direct effect was truncated from its original value of 381%.

To estimate direct effects and spillover effects, we used hierarchical targeted maximum likelihood estimation, a doubly-robust, semiparametric method that adjusts for potential confounders using ensemble machine learning (27). This approach is appropriate for cluster-level interventions that may result in statistical dependence between outcomes (see Methods) (28–30). We adjusted for covariates such as baseline malaria incidence and population size to account for differences in baseline characteristics between study arms (Tables S2-3).

In analyses of direct effects among intervention recipients, the chemoprevention intervention reduced malaria incidence among intervention recipients within 500m of index cases, but the confidence interval included the null. There was no evidence of a direct effect for the vector control or combined interventions, but precision was low (Figure 2, Table S4).

In analyses of spillover effects among intervention non-recipients up to 1 km away from interventions, chemoprevention and vector control interventions reduced incidence, but confidence intervals included the null. There was evidence of a spillover effect of the combined intervention, which reduced malaria incidence by 43% (95% CI 21%, 58%) among non-recipients.

We evaluated spillover effect heterogeneity by cluster-level malaria incidence and IRS coverage prior to the trial, surface temperature, rainfall, the enhanced vegetative index, elevation, and cohort-level treatment coverage, and gender. Across interventions, spillover effects were consistently more protective when pre-trial incidence was below the median (Figure S2). For example, the combined intervention reduced incidence by 68% (95% CI 35%, 84%) when baseline incidence was lower, but there was no effect when baseline incidence was above the median. Intervention spillover effects were generally stronger when environmental conditions favored mosquito breeding and survival (higher rainfall, higher enhanced vegetative index, and lower elevation) (Table S5). Spillover effects of the chemoprevention intervention were present for men but were null for women.

We performed several sensitivity analyses. When we repeated spillover effect analyses using 2 and 3 km radii around index cases to capture mosquito dispersal over longer distances (31, 32), results were similar (Figure S3). When using a shorter follow-up period in which intervention effects may have been stronger (see Methods), results were similar for the chemoprevention and vector control interventions; for the combined intervention, the spillover effect estimate was closer to the null, and precision was lower (Figure S4). When we estimated direct effects including the <3% of intervention recipients who resided >500m from index cases, results were nearly the same (Figure S5).

Overlap in analytic cohorts may have resulted in statistical dependence between outcomes (Figure 1; Table S6). When using alternative standard errors accounting for dependence (see Methods), confidence intervals were wider, but there was still evidence of spillover effects and total effects for the combined interventions (Table S4). When we excluded overlapping cohorts from the analysis, results were similar overall (Figure S4).

### Effects on malaria prevalence

We also estimated effects on malaria prevalence measured using qPCR in a cross-sectional survey at the end of the malaria season (May to August 2017). In contrast to incidence analyses, which captured any effects within the period immediately after intervention, prevalence analyses captured effects of cumulative interventions near the end of the trial. Prevalence also captures symptomatic and asymptomatic malaria cases that did not necessarily present at health clinics. Analyses of direct effects included individuals who resided within 500 m any intervention recipients; spillover effects included individuals with no intervention recipients < 500 m and at least one recipient within 500 m-3 km; total effects included individuals with at least one intervention recipient within 3 km (Figure 2b). We estimated prevalence ratios using targeted maximum likelihood estimation and adjusted standard errors for enumeration area-level clustering.

There was evidence of direct effects for all interventions, but confidence intervals included the null (Figure 2, Table S7). There was evidence of spillover effects: among non-recipients near intervention recipients, the chemoprevention intervention reduced prevalence by 72% (95% CI 31%, 88%), and the combined intervention reduced it by 79% (95% CI 6%, 95%). For the chemoprevention and combined interventions, spillover effects decreased in magnitude as distance to the nearest intervention increased (Figure 3). There was also evidence of spillover effects on prevalence of households with multiple malaria cases for the chemoprevention and combined interventions (Table S8).

**Figure 3.**
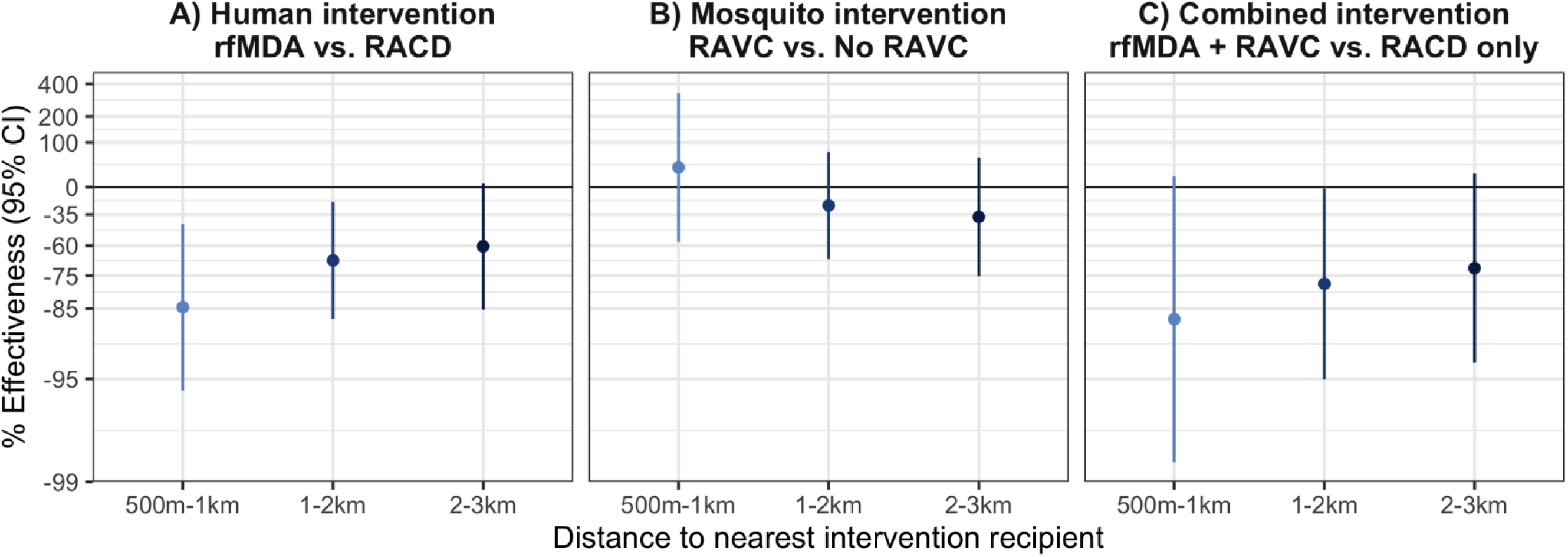
Spillover effects of reactive, focal malaria interventions on prevalence by distance to nearest intervention recipient. Spillover effects include individuals with no intervention recipients < 500m and any intervention recipients within different distance radii. **a**) Includes effects of the human intervention by comparing arms with rfMDA vs. those with RACD. **b**) Includes effects of the mosquito intervention by comparing arms with RAVC vs. those without RAVC. **c**) Includes effects of the combined intervention by comparing arms with rfMDA vs. RACD and rfMDA + RAVC vs. RACD only. Effects on prevalence estimated with TMLE using individual-level data; standard errors were adjusted for clustering at the enumeration area level. Models were adjusted for covariates that were screened separately for each model using a likelihood ratio test. Models were unadjusted because there were fewer than 10 malaria cases per variable. % Effectiveness was calculated as the ratio of incidence or prevalence between study arms minus 1 x 100%.

### Effects on malaria seroprevalence

We also investigated whether there were effects on seroprevalence of early transcribed membrane protein 5 antigen (Etramp5.Ag1), an indicator of recent malaria infection (33) that was measured by Luminex in the cross-sectional survey. The chemoprevention intervention reduced seroprevalence among individuals who resided within 500m of intervention recipients by 25% (95% CI 14%, 34%), and the combined intervention reduced it by 34% (95% CI 10%, 42%) (Figure 2, Table S9). Among intervention non-recipients, the combined intervention reduced seroprevalence by 34% (95% CI 20%, 45%).

### Cost-effectiveness

To inform policy decisions, we assessed cost-effectiveness using estimates of direct effects and spillover effects on prevalence. We calculated the incremental cost effectiveness ratio (ICER) by dividing the difference in cost between arms by the difference in prevalent cases averted between arms. We included cases averted for both individuals within 500m of any interventions and those with no intervention recipients < 500 m (direct effect population) and at least one recipient within 500 m-3 km (spillover effect population). Accounting for direct effects and spillover effects, the incremental cost-effectiveness ratios were $156 (95% CI $141, $177), $2,105 (95% CI $1,859, $2,430), and $1,142 (95% CI $944, $1,446) for the chemoprevention, vector control, and combined interventions (Table S10). Compared to the trial’s original incremental cost-effectiveness ratios estimates, accounting for spillover effects increased cost-effectiveness by 3%, 21%, and 37% for the chemoprevention, vector control, and combined interventions (34).

## Discussion

Here, we showed that a combined intervention of reactive focal chemoprevention plus IRS reduced malaria infections in intervention recipients as well as non-recipients up to 3 km away. Overall, spillover effects among non-recipients were strongest for the combined intervention, which was designed to reduce the parasite reservoir in both humans and mosquitoes. When accounting for spillover effects, the cost-effectiveness of the combined intervention was 37% higher than the prior estimate (34).

Interventions that produce spillover effects yield greater population health benefits at no additional cost. A prior analysis found that the combined intervention was highly cost-effective, but it did not account for possible spillover effects (34). When accounting for spillover effects, interventions were 3-37% more cost-effective (34). Given that malaria elimination requires substantially larger investments than malaria control (3, 35), evidence about cost savings due to spillover effects is critical to policy decisions about elimination strategies.

We found stronger evidence of spillover effects of the combined chemoprevention and vector control intervention over larger spatial scales than two prior studies of targeted malaria interventions. In Kenya, a trial in a low-transmission area found no change in parasite prevalence within 500m of serologically-defined hot spots that received targeted larviciding, long-lasting insecticide treated nets, IRS, and focal mass drug administration (19). In Zambia, an observational study in a high transmission setting found that IRS targeted to subdistricts with higher malaria incidence and population density reduced parasite prevalence in sprayed and unsprayed households within target areas; it did not measure spillover effects outside of target areas (20). The interventions in our study may have been more likely to produce spillover effects because they were delivered repeatedly in response to subsequent index cases. In this trial, clusters received interventions up to 17 times per cluster; they were repeated annually over three years in the Zambia study and once in the Kenya trial. Further, it is possible that delivering interventions in response to new index cases can more effectively reduce transmission than targeting interventions based on an area’s incidence or seroprevalence.

For the chemoprevention intervention, there was evidence of spillover effects on prevalence and suggestive evidence of spillover effects and direct effects on incidence. Incidence analyses measured effects shortly after interventions, while prevalence analyses measured them at the end of the transmission season. Thus, our findings may indicate that reductions in local transmission accrued as additional rounds of chemoprevention interventions were delivered and population intervention coverage increased. This may especially be the case for the chemoprevention intervention since reductions in infectiousness of malaria cases are typically short-lived following treatment, especially in the absence of concurrent vector control (36). Overall, these findings suggest that reactive, focal chemoprevention can more effectively reduce asymptomatic or subclinical infections among nearby non-intervention recipients than RACD, particularly after repeated rounds.

For the vector control intervention, the primary analysis did not find direct effects, and the spillover effect estimate included the null despite strong biologic plausibility for both types of effects. We used a 6-month follow-up period to capture longer-term effects of IRS, which resulted in spatiotemporal overlap between analytic cohorts (Table S6). This overlap may have induced dependence between outcomes that was not fully accounted for by covariate adjustment, resulting in residual bias (28). Analyses of current infection prevalence were not subject to concerns about cohort overlap and were suggestive of direct effects, but confidence intervals included the null, and there was no evidence of spillover effects. Finally, our pre-specified subgroup analyses suggested that spillover effects of RAVC were driven by baseline transmission levels and environmental conditions: spillover effects on incidence were present in areas with baseline malaria incidence was <14 per 1,000 and when weather conditions favored mosquito breeding and survival (temperature < 31 °C; monthly rainfall < 27 mm).

The combined intervention appeared to have synergist effects, reducing local transmission to intervention non-recipients via spillover effects in all analyses. This may be because short-lived reductions in host infectiousness following chemotherapy can be sustained over time when coupled with IRS’ long-lasting reductions in mosquito populations. In effect, each intervention reduces the parasite reservoirs in hosts and vectors, and the combination of interventions slows the replenishment of parasite reservoirs (36). This may explain why we found that spillover effects were larger for prevalence of current infection, which captured effects at the end of the season, rather than incidence, which captured short-term effects. Our findings are consistent with two recent studies that found evidence of potential synergistic community-level effects when combining community-wide chemoprevention or seasonal malaria chemoprevention with IRS in high transmission settings (37, 38). Results are also consistent with a modeling study that estimated that the joint effect of chemoprevention and IRS was over 1.5 times higher than effects of intervention alone in low-transmission settings (36). Taken together, our results suggest that the combined intervention may be particularly effective as a reactive intervention or outbreak response in low-transmission settings approaching elimination or possibly following introduction of cases after elimination has been achieved.

Our estimates of direct and spillover effects shed light on the mechanism through which these targeted interventions work in time and space. We found that spillover effect sizes were generally similar to or stronger than direct effect sizes. It is possible that during the time between index case detection and intervention delivery (median 13-14 days) (24), transmission occurred to nearby intervention recipients. Thus, the interventions may not have been rapid enough to reduce malaria among recipients but may have prevented onward transmission to non-recipients further from index cases. In addition, our finding that spillover effects on prevalence were stronger at shorter distances to interventions suggests that the majority of the spillover effect occurred within 1km of index cases.

Our study was subject to several limitations. First, due to rare outcomes, precision was low in some analyses and may have increased the chance of Type II error, especially for direct effects. Second, when constructing analytic cohorts, household relocation after baseline could have resulted in misclassification of households to target areas or spillover zones. Third, incidence analyses could not fully rely on randomization-based inference due to cohort overlap; it is possible that covariate adjustment did not fully account for imbalances between arms. In future studies, using a ring trial design to test focal interventions could improve baseline balance, increase precision, and minimize overlap between target areas (39).

Despite these limitations, the internal consistency between the findings of this secondary analysis and the original trial, which each used different data structures and statistical methods, supports the validity of our findings. Estimates of total effects in this analysis, which pooled across intervention recipients and non-recipients, were consistent overall with those of the original trial, which included all individuals in study clusters (intervention recipients and non-recipients) (24, 40). Additional strengths include pre-specification of spillover analysis methods and use of individual-level, spatially indexed data to measure spillovers.

In conclusion, we found that reactive, focal malaria interventions targeting both human and mosquito parasite reservoirs reduced malaria risk, even among non-intervention recipients up to 3km from index cases. Further, the combined intervention could be particularly useful in responding to imported infections, which pose a persistent threat prior to and following elimination. Our findings suggest that these interventions are an effective strategy for achieving and maintaining malaria elimination.

## Materials and Methods

### Analysis overview

This study was a secondary analysis of a cluster-randomized trial of focal malaria interventions conducted in Zambezi region of Namibia from January 1 to December 31, 2017 (NCT02610400) (41, 24). Using a two-by-two factorial design, the trial randomly allocated 56 study clusters to study arms: reactive case detection (RACD) only, reactive focal mass drug administration (rfMDA) only, RACD + reactive vector control (RAVC), and rfMDA + RAVC. Here, we separately estimated effects among intervention recipients and non-recipients to estimate direct effects and spillover effects. We estimated the effects of the chemoprevention intervention (rfMDA vs. RACD), the vector control intervention (RAVC vs. no RAVC), or combined interventions (rfMDA + RAVC vs. RACD only), consistent with the original trial (24). The analysis plan for this study was pre-specified at https://osf.io/s8ay4/. Deviations from the pre-analysis plan and details about the study site and trial are in the Supporting Information.

### Ethics statement

The trial protocol was approved by the Namibia Ministry of Health and Social Services (17/3/3) and the Institutional Review Boards at the University of California San Francisco (15–17422) and London School of Hygiene & Tropical Medicine (10411). The secondary analysis protocol was approved by the Stanford University Institutional Review Board (60708).

### Incidence analyses

The unit of intervention (index cases) and the unit of randomization (clusters) differed, so cluster-level analyses would not have captured fine-scale direct effects and spillover effects. To capture the person-time in which we expected each intervention to influence incident malaria infections, we created analytic cohorts in space and time around each index case that triggered an intervention. The primary analysis used a 1 km radius around each index case because the majority of mosquito movement occurs within < 1 km.

We pre-specified cohort follow-up length based on the period in which we expected each intervention to reduce malaria among intervention recipients (direct effects) and non-recipients (spillover effects). For comparisons of rfMDA and RACD interventions, the direct effect follow-up period was 35 days, the length of intrinsic incubation period for *Pf* malaria (8). The spillover effect follow-up period was 21 to 56 days; the 3-week lag period allowed for gametocyte clearance in the treated individual, sporozoite development in mosquitos, and development of detectable merozoites in humans. For RAVC interventions, the direct effects follow-up period was 6 months since IRS can remain effective for an entire transmission season (9). The spillover effects follow-up period was from day 17 to 6 months. We conducted a sensitivity analysis with alternative follow-up lengths (rfMDA and RACD direct effects: day 0-21; spillover effects: day 21-42; RAVC direct effects day 0-7; spillover effects day 17-90). Additional details are in the Supporting Information.

### Statistical models for incidence

To compare incidence between arms, we used hierarchical targeted maximum likelihood estimation (TMLE), a double robust, semi-parametric approach appropriate for cluster-level exposures (27). TMLE estimates both an outcome regression and a propensity score (the probability of treatment conditional on covariates) and updates the initial parameter estimate using information in the propensity score. Compared to other parametric models for clustered data (e.g., mixed effects models, generalized estimating equations), hierarchical TMLE imposes fewer assumptions and may be more efficient for randomized trials (42). We fit outcome and propensity score models using an ensemble machine learning algorithm (the Superlearner) (43). We adjusted standard errors to account for potential correlation due to overlap between some cohorts using a model of cohort-level influence curves analogous to variance-covariance models used in cross-random effects models (See details in Supporting Information) (44, 45).

Because incidence analyses did not rely on cluster randomization, we adjusted for covariates that were correlated with the outcome (likelihood ratio test p-value < 0.2) (46). Propensity score models adjusted for the following baseline variables: cluster-level indoor residual spray coverage, malaria incidence, median monthly rainfall, median enhanced vegetative index, and median daytime land surface temperature in the season prior to the trial, population size, and median elevation. Outcome models adjusted for individual- and cluster-level covariates.

Individual-level covariates included sex, age, calendar month of intervention, distance from an individual’s residence to the residence of the index case that triggered an intervention, number of interventions an individual previously received, number of previously intervention recipients within 0.5, 1, 2, and 3 km of the individual’s residence (from the start of the trial to the start of the cohort’s observation period), and population size within 0.5, 1, 2, and 3 km of the individual’s residence. Cluster-level covariates included those in the propensity score models as well as mean distance to the nearest neighboring household, mean distance to the nearest healthcare facility, and mean time from index case detection to intervention.

### Prevalence analyses

To capture effects of cumulative interventions at the end of the malaria transmission season, we estimated effects on prevalence using data from a cross-sectional survey. Direct effects analyses included individuals who resided near any intervention recipients within 500 m of their residence, spillover effects analyses included individuals with no intervention recipients < 500 m and at least one recipient within 500 m-3 km, and total effects included individuals with at least one intervention recipient within 3 km.

### Statistical models for prevalence

Prevalence analyses used data from the cross-sectional survey conducted at the end of the malaria transmission season in 2017. We used targeted maximum likelihood estimation (47) with individual-level data with the same learners included in incidence analyses. We accounted for correlation within enumeration area-level clusters using cluster-level influence curve-based standard errors. The covariate set included the same cluster-level covariates included in incidence analyses and the following individual-level covariates: age, sex, occupation, recent travel, household slept under a bed net, slept outdoors in the past two weeks, and the total population, number of intervention recipients, number of intervention recipients in the same study arm, the number of intervention recipients in a different study arm, and the proportion of intervention recipients with the same treatment within 500 m, 1 km, 2 km, and 3 km of sampled individuals. We screened for covariates using the same approach described for incidence analyses.

### Cost-effectiveness analysis

We estimated incremental cost effectiveness ratios (ICER) incorporating direct effects and spillover effects. To facilitate comparison with the original cost-effectiveness estimates for the trial (34), we used estimated effects on prevalence measured by qPCR. ICERs estimated using incidence would not be directly comparable between this study and the original trial because we used a cohort-level analysis, but the original trial used a cluster-level analysis. We used previously published estimates of total intervention costs in 2017 in US dollars (34). To obtain the number of prevalent cases averted, we multiplied the difference in prevalence between arms among intervention recipients and non-recipients by the estimated population size included in direct effects and spillover effects analyses. We calculated the ICER by dividing the difference in cost between arms by the difference in prevalent cases averted between arms among individuals who were located within 500m of any intervention recipients and individuals who resided within 500m to 3 km of interventions.

## Data and Code Availability

Data from the original trial is available at https://clinepidb.org/ce/app/workspace/analyses/DS_f559aee789. Replication scripts are available at https://github.com/jadebc/malaria-focal-spillover-public.

## Author Contributions

JBC and MH conceptualized the study. JBC, AB, AH, MVDL, and MH developed the analysis plan. HN, LMP, MT, LW, CD, RG, DM, IK, and MH contributed data. JBC, HL, AN, and GBH processed data. JBC and HL conducted the analysis. JBC wrote the manuscript, and all authors edited the manuscript.

## Competing Interest Statement

The authors have no competing interests to declare.

## Acknowledgements

The original trial was supported by Novartis Foundation (A122666), the Bill & Melinda Gates Foundation (OPP1160129), and the Horchow Family Fund (5300375400). Research reported in this publication was supported in part by the National Institute of Allergy and Infectious Diseases of the National Institutes of Health under Award Numbers K01AI141616 (PI: Benjamin-Chung) and the National Heart, Lung, And Blood Institute of the National Institutes of Health under award number T32HL151323 (Nguyen) and Novartis Foundation Supplement to A122666 (PIs: Gosling, Hsiang). The content is solely the responsibility of the authors and does not necessarily represent the official views of the National Institutes of Health. Jade Benjamin-Chung and Michelle Hsiang are Chan Zuckerberg Biohub Investigators. We also acknowledge the Stanford Research Computing Center for computational resources at the Sherlock high-performance cluster.

---

**Figure S1.**
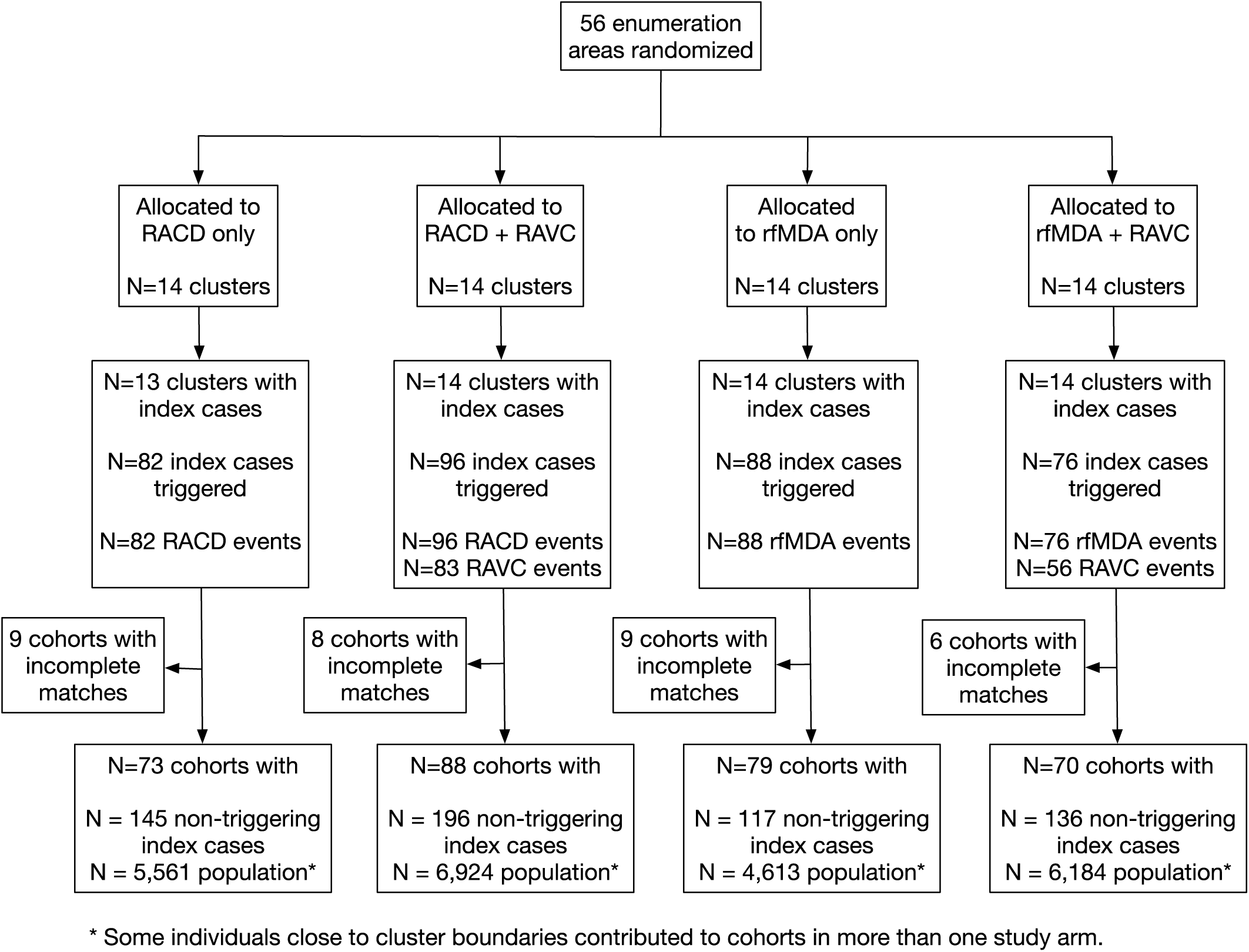
Diagram of study randomization, index cases, and population by arm. RACD: reactive case detection. rfMDA: reactive, focal mass drug administration. RAVC: reactive vector control.

**Figure S2.**
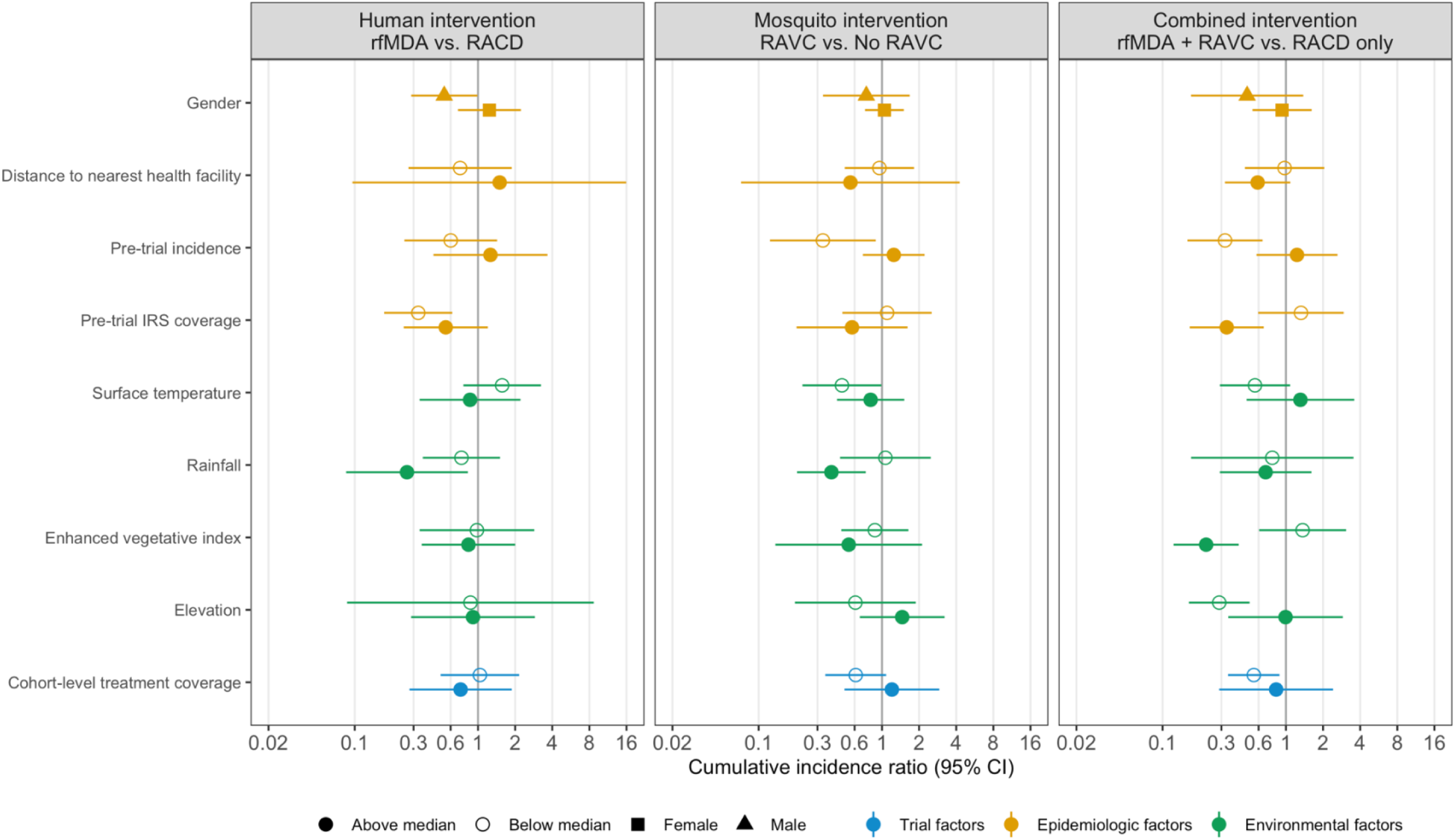
Spillover effect estimates on cumulative incidence within subgroups. Cumulative incidence ratios estimated with hierarchical TMLE; outcome models were fit with cohort-level data. Models were adjusted for covariates that were screened separately for each model using a likelihood ratio test. Models for rfMDA + RAVC vs. RACD were unadjusted due to data sparsity. Confidence intervals account for cohort overlap. For rfMDA and RACD arms, the analysis includes the period from 0-35 days following index case detection for direct effects and 21-56 days for spillover effects. For rfMDA+RAVC and RAVC only arms, the analysis includes the period from 0-6 months following index case detection for direct effects and 17 days to 6 months for spillover effects. Total effects analyses include the person-time for the direct effects and spillover effects analyses. Direct effect includes treated in target zone. Spillover effect includes intervention non-recipients up to 1km from an index case. Total effect includes all individuals (intervention recipients and non-recipients) up to 1km from index case. For the human intervention, confidence interval upper bounds were truncated at 16 for above median distance to the nearest health facility (observed value: 23).

**Figure S3.**
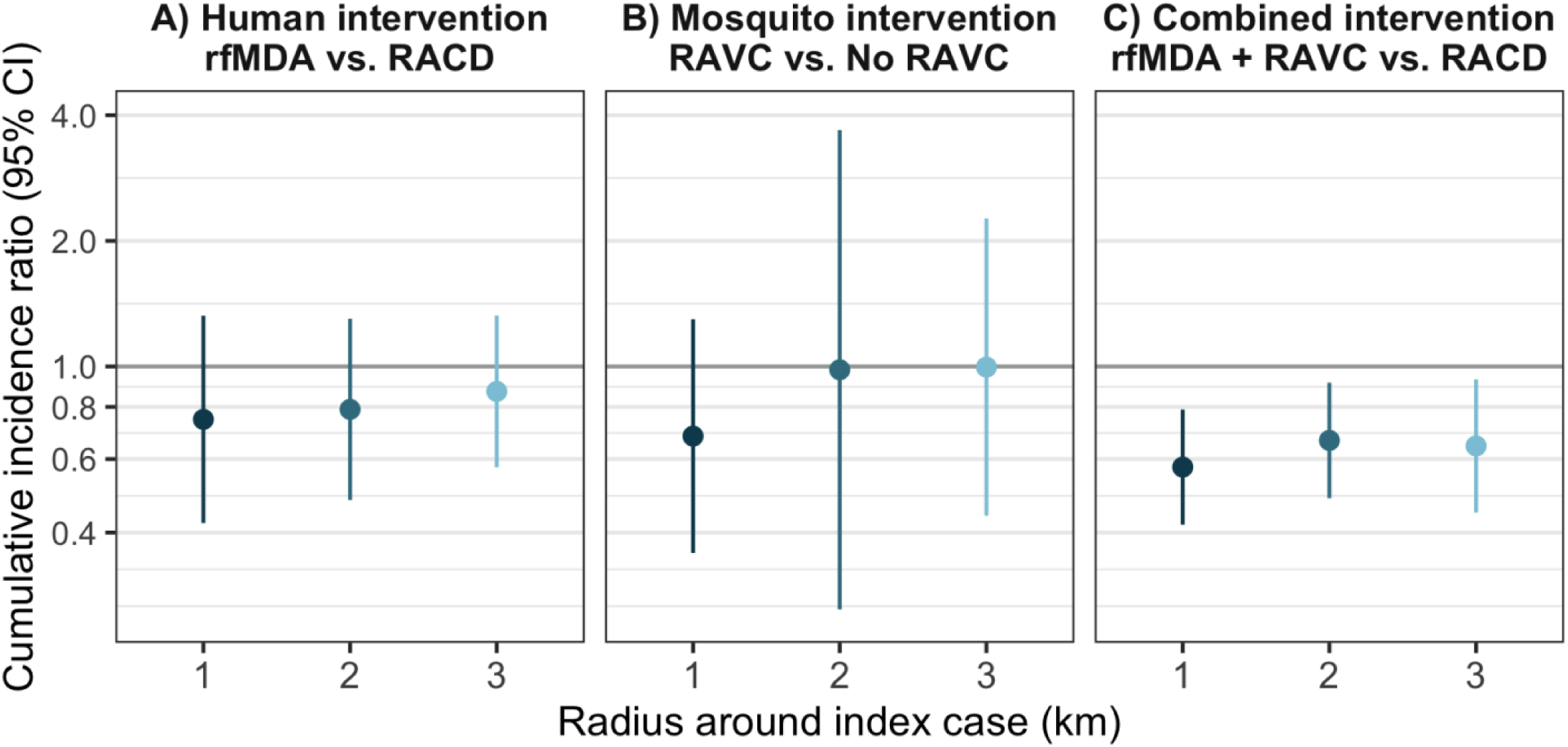
Sensitivity analyses for spillover effects on cumulative incidence of malaria with different distance radii. For rfMDA and RACD arms, the primary analysis includes the period from 0-35 days following index case detection for direct effects and 21-56 days for spillover effects; the alternative observation period analysis includes the period from 0-21 days following index case detection for direct effects and 21 to 42 days for spillover effects. For rfMDA+RAVC and RAVC only arms, the primary analysis includes the period from 0-6 months following index case detection for direct effects and 17 days to 6 months for spillover effects; the alternative observation period analysis includes the period from 0-7 days following index case detection for direct effects and 17 to 90 days for spillover effects. Total effects analyses include the person-time for the direct effects and spillover effects analyses. Direct effect includes intervention recipients in target zone. Spillover effect includes intervention non-recipients up to 1km from an index case in the primary analysis and up to 2km or 3km in sensitivity analyses. Total effect includes all individuals (intervention recipients and non-recipients) up to 1km from index case in the primary analysis and up to 2km or 3km in sensitivity analyses. Includes cohort-level analyses for all estimates except spillover effects of the combined intervention. All incidence outcome models were fit with cohort-level data except for models of spillover effects of rfMDA vs. RACD and rfMDA + RAVC vs. RACD only.

**Figure S4.**
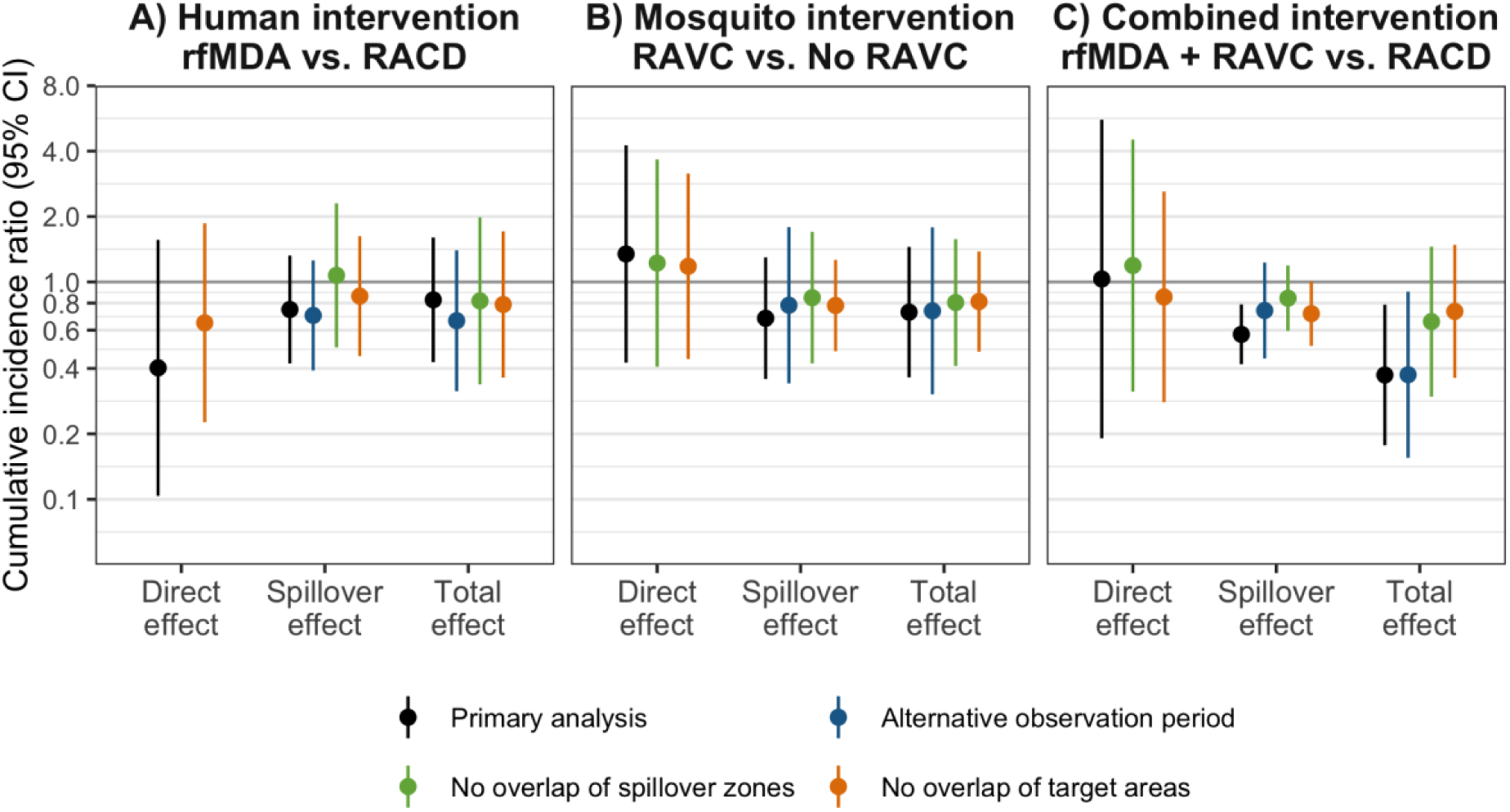
Sensitivity analyses for effects on cumulative incidence of malaria. For rfMDA and RACD arms, the primary analysis includes the period from 0-35 days following index case detection for direct effects and 21-56 days for spillover effects; the alternative observation period analysis includes the period from 0-21 days following index case detection for direct effects and 21 to 42 days for spillover effects. For rfMDA+RAVC and RAVC only arms, the primary analysis includes the period from 0-6 months following index case detection for direct effects and 17 days to 6 months for spillover effects; the alternative observation period analysis includes the period from 0-7 days following index case detection for direct effects and 17 to 90 days for spillover effects. Total effects analyses include the person-time for the direct effects and spillover effects analyses. Direct effect includes intervention recipients in target zone. Spillover effect includes intervention non-recipients up to 1km from an index case. Total effect includes all individuals (intervention recipients and non-recipients) up to 1km from index case. Sensitivity analyses for no overlap of spillover zones excluded any cohorts whose spillover zones overlapped spatially or temporally with other spillover zones. Sensitivity analyses for no overlap of target areas excluded any cohorts whose target areas overlapped spatially or temporally with other target areas. Some direct effects models could not be fit due to data sparsity. All incidence outcome models were fit with cohort-level data except for models of spillover effects of rfMDA vs. RACD and rfMDA + RAVC vs. RACD only.

**Figure S5.**
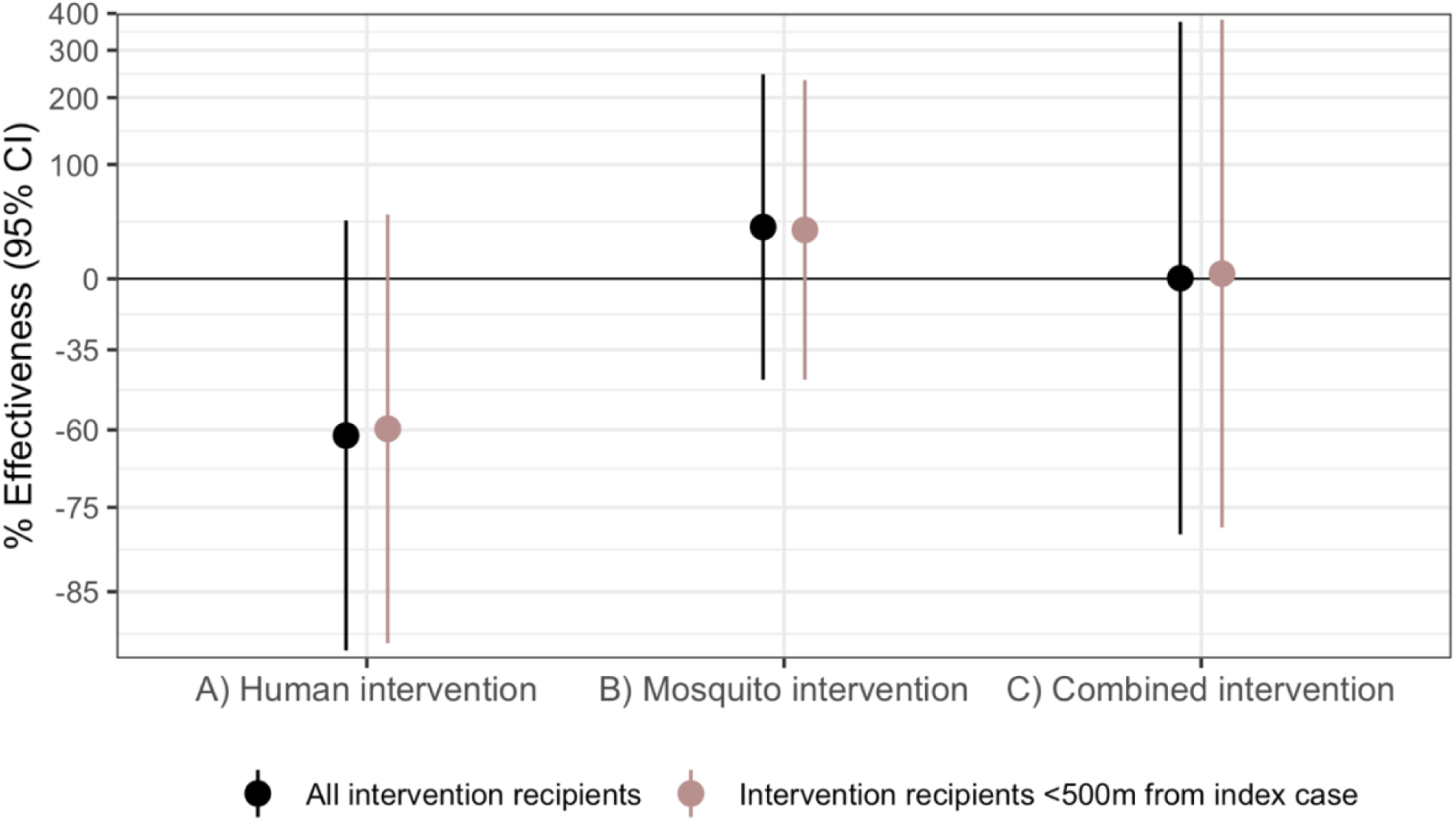
Sensitivity analyses for direct effects including all intervention recipients. The observation period was 0-35 days for rfMDA and RACD arms and 0-6 months for rfMDA+RAVC and RAVC only arms. Black points indicate estimates from analyses including all intervention recipients, regardless of whether they resided within the target zone within 500m of index cases. Mauve points indicate estimates from analyses restricting to intervention recipients within 500m of index cases that triggered interventions. Analyses were performed at the cohort level.

**Table S1.** Two-by-two factorial study design of reactive focal interventions. Reactive case detection (RACD) involved administering rapid diagnostic tests for malaria to individuals living within a 500-m radius of an index case and treating individuals who tested positive with artemether-lumefantrine and single-dose primaquine. Reactive focal mass drug administration (rfMDA) involved presumptively treating individuals living within a 500-m radius of an index case with artemether-lumefantrine, without testing for malaria beforehand. Reactive focal vector control (RAVC) involved spraying the long-lasting insecticide, pirimiphos-methyl, to the interior walls of households located within a seven-household radius of an index case. The effectiveness of three interventions were compared to three respective controls: (1) rfMDA versus RACD (B and D vs A and C); (2) RAVC versus no RAVC (C and D vs A and B); and (3) rfMDA plus RAVC versus a RACD only (D vs A). Reproduced from Hsiang et al. 2020 *Lancet* with permission.

|  |  | Human intervention |  |
| --- | --- | --- | --- |
|  |  | Reactive case detection only<br>(28 clusters) | Reactive focal mass drug administration<br>(28 clusters) |
| Mosquito intervention | No reactive focal vector control<br>(28 clusters) | Reactive case detection only<br>(14 clusters) | Reactive focal mass drug administration only<br>(14 clusters) |
|  | Reactive focal vector control<br>(28 clusters) | Reactive case detection plus reactive focal vector control<br>(14 clusters) | Reactive focal mass drug administration plus reactive focal vector control<br>(14 clusters) |

**Table S2.** Baseline characteristics among intervention recipients. Includes data from intervention recipients in target areas located within 500m of an index case.

|  | Human intervention |  | Mosquito intervention |  | Human & mosquito intervention |  |
| --- | --- | --- | --- | --- | --- | --- |
|  | RACD | rfMDA | No RAVC | RAVC | RACD only | rfMDA + RAVC |
| **Population characteristics** |  |  |  |  |  |  |
| Number of cohorts | 161 | 149 | 152 | 158 | 73 | 70 |
| Mean cohort population size (SE) | 26 (1) | 27 (1) | 26 (1) | 27 (1) | 26 (1) | 29 (1) |
| Mean cluster population size (SE) | 389.6 (1.94) | 346.4 (1.96) | 358.9 (2.01) | 376.9 (1.94) | 353.0 (2.05) | 328.0 (1.89) |
| Malaria incidence per 1,000 in 2016 (SE) | 27.0 (0.37) | 55.8 (1.26) | 31.9 (0.60) | 50.0 (1.15) | 26.6 (0.56) | 75.4 (2.25) |
| Pre-season indoor residual spray coverage 2016 (SE) | 76.3 (0.32) | 77.1 (0.36) | 77.9 (0.37) | 75.6 (0.31) | 83.6 (0.43) | 81.5 (0.42) |
| Distance to nearest healthcare facility (km) (SE) | 5.2 (0.06) | 6.7 (0.08) | 5.0 (0.06) | 6.7 (0.07) | 3.5 (0.06) | 6.9 (0.12) |
| **Ecological factors (range)** |  |  |  |  |  |  |
| Median monthly rainfall November 2016-April 2017 (mm) | 23.7 (18.4, 26.7) | 23.5 (18.4, 26.7) | 23.5 (18.4, 26.7) | 23.7 (18.4, 26.7) | 23.7 (18.4, 26.7) | 23.7 (18.4, 26.7) |
| Median enhanced vegetative index January 2017-July 2017 | 0.15 (0.09, 0.31) | 0.15 (0.09, 0.27) | 0.15 (0.09, 0.22) | 0.15 (0.09, 0.31) | 0.15 (0.10, 0.21) | 0.15 (0.09, 0.27) |
| Median elevation (m) | 522 (387, 1021) | 541 (412, 1124) | 527 (398, 1124) | 547 (387, 1021) | 522 (398, 921) | 576 (412, 984) |
| Median daytime land surface temperature (C) | 30.5 (28.9, 33.4) | 31.1 (28.6, 32.5) | 30.7 (28.6, 33.4) | 30.8 (28.7, 32.5) | 30.7 (28.9, 33.4) | 31.1 (28.7, 32.5) |

**Table S3.**
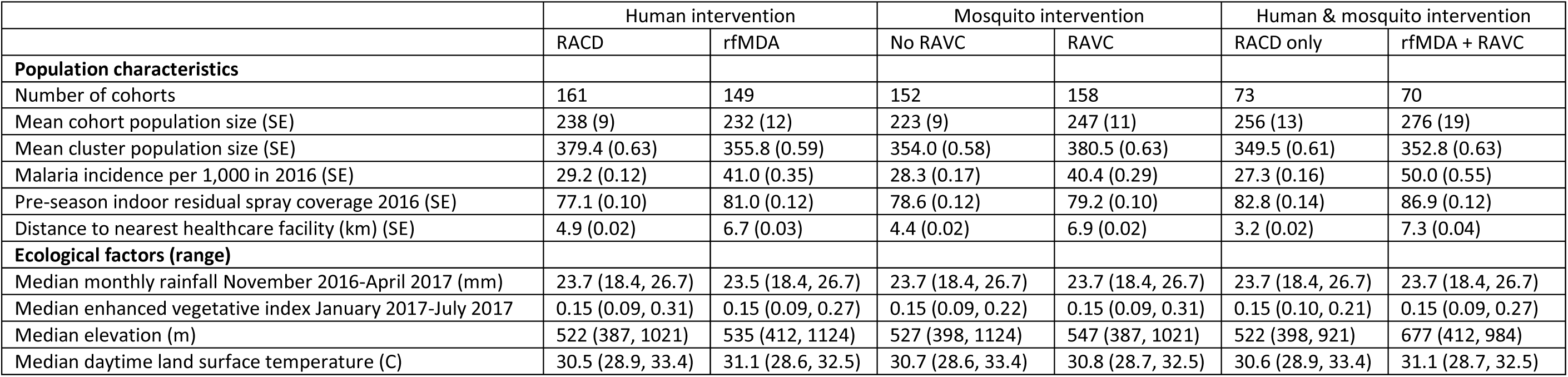
Baseline characteristics among non-intervention recipients up to 1km away from index cases. Includes data from intervention non-recipients up to 1km from an index case that triggered interventions.

**Table S4.** Direct effect, spillover effect, and total effect estimates on cumulative incidence of malaria infection. For rfMDA and RACD arms, the analysis includes the period from 0-35 days following index case detection for direct effects and 21-56 days for spillover effects. For rfMDA+RAVC and RAVC only arms, the analysis includes the period from 0-6 months following index case detection for direct effects and 17 days to 6 months for spillover effects. Total effects analyses include the person-time for the direct effects and spillover effects analyses. Direct effect includes intervention recipients in the target zone. Spillover effect analyses includes intervention non-recipients up to 1km from an index case. Total effect includes all individuals (intervention recipients and non-recipients) up to 1km from index case. Models were fit with hierarchical targeted maximum likelihood. All outcome models were fit with cohort-level data except for models of spillover effects of rfMDA + RAVC vs. RACD only. Adjusted models were fit if there were fewer than 10 malaria cases per variable. Covariates were screened separately for each model using a likelihood ratio test. We separately fit individual- and cohort-level outcome models and report the model with the smaller cross-validated mean squared error. All models except spillover effects of the human and combined interventions were fit on cohort-level data.

|  |  |  | Incidence proportion |  | Incidence ratio (95% CI) |  |  |
| --- | --- | --- | --- | --- | --- | --- | --- |
|  | N cohorts | N | Intervention arm | Reference arm | Unadjusted | Adjusted | Adjusted, CI adjusted for cohort overlap |
| **Human intervention (rfMDA vs. RACD)** |  |  |  |  |  |  |  |
| Direct effect | 310 | 8,252 | 3.4 | 6.5 | 0.53 (0.25, 1.11) | 0.40 (0.11, 1.48) | 0.40 (0.10, 1.56) |
| Spillover effect | 310 | 72,830 | 9.0 | 9.9 | 0.91 (0.60, 1.37) | 0.82 (0.52, 1.29) | 0.82 (0.44, 1.51) |
| Total effect | 310 | 81,082 | 8.4 | 9.6 | 0.88 (0.59, 1.31) | 0.83 (0.51, 1.35) | 0.83 (0.43, 1.60) |
| **Mosquito intervention (RAVC vs. no RAVC)** |  |  |  |  |  |  |  |
| Direct effect | 310 | 8,252 | 8.9 | 7.6 | 1.17 (0.62, 2.23) | 1.35 (0.54, 3.34) | 1.35 (0.43, 4.25) |
| Spillover effect | 310 | 72,830 | 12.9 | 18.5 | 0.69 (0.47, 1.03) | 0.68 (0.46, 1.00) | 0.68 (0.36, 1.30) |
| Total effect | 310 | 81,082 | 12.5 | 17.4 | 0.72 (0.49, 1.06) | 0.73 (0.49, 1.08) | 0.73 (0.36, 1.45) |
| **Combined intervention (rfMDA + RAVC vs. RACD only)** |  |  |  |  |  |  |  |
| Direct effect | 143 | 3,914 | 6.4 | 7.4 | 0.87 (0.32, 2.41) | 1.03 (0.22, 4.81) | 1.03 (0.19, 5.58) |
| Spillover effect | 143 | 38,048 | 11.2 | 18.1 | 0.62 (0.34, 1.13) | 0.57 (0.41, 0.80) | 0.57 (0.42, 0.79) |
| Total effect | 143 | 41,962 | 10.8 | 17.1 | 0.63 (0.35, 1.12) | 0.37 (0.22, 0.63) | 0.37 (0.18, 0.79) |

**Table S5.** Range above and below median value in each enumeration area for subgroup variables.

|  | Below median |  | Above median |  |
| --- | --- | --- | --- | --- |
|  | Minimum | Maximum | Minimum | Maximum |
| Malaria incidence per 1,000 in 2016 | 0.0 | 13.9 | 14.9 | 293.3 |
| Pre-season indoor residual spray coverage 2016 (%) | 27.2 | 77.3 | 77.9 | 100 |
| Median daytime land surface temperature (C) | 28.6 | 31.1 | 31.1 | 33.4 |
| Median monthly rainfall November 2016-April 2017 (mm) | 18.4 | 23.7 | 23.7 | 26.7 |
| Median enhanced vegetative index January 2017-July 2017 | 0.09 | 0.15 | 0.15 | 0.31 |
| Median elevation (m) | 387 | 541 | 544 | 1124 |
| Cohort-level treatment coverage (%) | 0.0 | 8.3 | 8.3 | 97.4 |

**Table S6.** Percentage of cohorts overlapping with other cohorts. Overlap in target area was defined as index cases that triggered interventions located within <1km of each other and observation periods that temporally overlapped with another cohort’s. Overlap in spillover zones was defined as index cases that triggered interventions located within 1-2km of each other and observation periods that temporally overlapped with another cohort’s. The denominator was the total cohorts included in each analysis.

|  | Primary analysis |  | Sensitivity analysis with shorter observation period |  |
| --- | --- | --- | --- | --- |
|  | Target areas | Spillover zone | Target areas | Spillover zone |
| **Human intervention**<br>(rfMDA vs. RACD) | 32.0 | 28.9 | 21.2 | 18.4 |
| **Mosquito intervention**<br>(RAVC vs. no RAVC) | 59.2 | 47.5 | 53.8 | 41.8 |
| **Combined intervention**<br>(rfMDA + RAVC vs. RACD only) | 60.5 | 28.1 | 60.2 | 24.1 |

**Table S7.** Direct effect, spillover effect, and total effect estimates on malaria prevalence measured by qPCR. Prevalence was measured in a cross-sectional survey in a random sample of households at the end of the malaria season. Analyses were restricted to individuals located within 3 km of at least one intervention recipient. Direct effects include individuals with any intervention recipients within 500m, spillover effects include individuals with no intervention recipients < 500m and any intervention recipients 500m-3km, and total effects include individuals with any intervention recipients <3km during the study. Prevalence ratios were estimated using TMLE with individual-level data, and standard errors were adjusted for clustering at the enumeration area level. Adjusted analyses were not fit there were fewer than 30 observations within strata of the intervention and outcome. Adjusted models were not fit if the number of cases within treatment arm strata was <30.

|  | N |  | Prevalence |  | Prevalence ratio (95% CI) |  |
| --- | --- | --- | --- | --- | --- | --- |
|  | Intervention arm | Reference arm | Intervention arm | Reference arm | Unadjusted | Adjusted |
| **Human intervention**<br>(rfMDA vs. RACD) |  |  |  |  |  |  |
| Direct effect | 1537 | 1835 | 0.029 | 0.033 | 0.90 (0.61, 1.31) | 0.84 (0.53, 1.32) |
| Spillover effect | 244 | 229 | 0.025 | 0.087 | 0.28 (0.12, 0.69) | -- |
| Total effect | 1781 | 2064 | 0.029 | 0.039 | 0.74 (0.52, 1.04) | 0.79 (0.51, 1.19) |
| **Mosquito intervention**<br>(RAVC vs. no RAVC) |  |  |  |  |  |  |
| Direct effect | 1710 | 1662 | 0.026 | 0.037 | 0.70 (0.48, 1.03) | 0.78 (0.51, 1.21) |
| Spillover effect | 195 | 278 | 0.051 | 0.058 | 0.89 (0.41, 1.92) | -- |
| Total effect | 1905 | 1940 | 0.028 | 0.040 | 0.71 (0.51, 1.01) | 0.64 (0.43, 0.96) |
| **Combined intervention**<br>(rfMDA + RAVC vs. RACD only) |  |  |  |  |  |  |
| Direct effect | 758 | 883 | 0.017 | 0.033 | 0.52 (0.27, 1.00) | -- |
| Spillover effect | 118 | 152 | 0.017 | 0.079 | 0.21 (0.05, 0.94) | -- |
| Total effect | 876 | 1035 | 0.017 | 0.040 | 0.43 (0.24, 0.78) | -- |

**Table S8.** Direct effect, spillover effect, and total effect estimates on household-level malaria prevalence of measured by qPCR. Prevalence was measured in a cross-sectional survey in a random sample of households at the end of the malaria season. Analyses were run at the household level. Household-level malaria prevalence was the percentage of households with more than one malaria case detected in the prevalence survey by qPCR. Direct effects include households with any intervention recipients within 500m, spillover effects include households with no intervention recipients < 500m and any intervention recipients 500m-3km, and total effects include households with any intervention recipients <3km during the study. Prevalence ratios were estimated using TMLE with household-level data. Adjusted analyses were not fit there were fewer than 30 observations within strata of the intervention and outcome. Adjusted models were not fit if the number of cases within treatment arm strata was <30.

|  | N households |  | Prevalence |  | Unadjusted<br>Prevalence Ratio<br>(95% CI) |
| --- | --- | --- | --- | --- | --- |
|  | Intervention arm | Reference arm | Intervention arm | Reference arm |  |
| **Human intervention**<br>(rfMDA vs. RACD) |  |  |  |  |  |
| Direct effect | 456 | 506 | 0.018 | 0.018 | 0.99 (0.38, 2.54) |
| Spillover effect | 72 | 69 | 0.000 | 0.043 | 0.00 (0.00, 0.00) |
| Total effect | 528 | 575 | 0.015 | 0.021 | 0.73 (0.30, 1.76) |
| **Mosquito intervention**<br>(RAVC vs. no RAVC) |  |  |  |  |  |
| Direct effect | 481 | 481 | 0.012 | 0.023 | 0.55 (0.20, 1.46) |
| Spillover effect | 65 | 76 | 0.015 | 0.026 | 0.58 (0.05, 6.35) |
| Total effect | 546 | 557 | 0.013 | 0.023 | 0.55 (0.22, 1.37) |
| **Combined intervention**<br>(rfMDA + RAVC vs. RACD only) |  |  |  |  |  |
| Direct effect | 219 | 244 | 0.005 | 0.016 | 0.28 (0.03, 2.48) |
| Spillover effect | 36 | 40 | 0.000 | 0.050 | 0.00 (0.00, 0.00) |
| Total effect | 255 | 284 | 0.004 | 0.021 | 0.19 (0.02, 1.53) |

**Table S9.** Direct effect, spillover effect, and total effect estimates on Etramp5.Ag1 seroprevalence. Prevalence was measured in a cross-sectional survey in a random sample of households at the end of the malaria season. Analyses were restricted to individuals located within 3 km of at least one intervention recipient. Direct effects include individuals with any intervention recipients within 500m, spillover effects include individuals with no intervention recipients < 500m and any intervention recipients 500m-3km, and total effects include individuals with any intervention recipients <3km during the study. Prevalence ratios were estimated using TMLE with individual-level data, and standard errors were adjusted for clustering at the enumeration area level. Adjusted analyses were not fit there were fewer than 30 observations within strata of the intervention and outcome. Adjusted models were not fit if the number of cases within treatment arm strata was <30.

|  | N |  | Prevalence |  | Prevalence ratio (95% CI) |  |
| --- | --- | --- | --- | --- | --- | --- |
|  | Intervention arm | Reference arm | Intervention arm | Reference arm | Unadjusted | Adjusted |
| **Human intervention**<br>(rfMDA vs. RACD) |  |  |  |  |  |  |
| Direct effect | 1316 | 1611 | 0.215 | 0.285 | 0.75 (0.66, 0.86) | 0.84 (0.71, 1.00) |
| Spillover effect | 198 | 182 | 0.227 | 0.225 | 1.01 (0.69, 1.46) | 1.32 (0.73, 2.41) |
| Total effect | 1514 | 1793 | 0.217 | 0.279 | 0.78 (0.69, 0.88) | 0.85 (0.73, 0.99) |
| **Mosquito intervention**<br>(RAVC vs. no RAVC) |  |  |  |  |  |  |
| Direct effect | 1475 | 1452 | 0.241 | 0.267 | 0.90 (0.80, 1.02) | 0.90 (0.79, 1.04) |
| Spillover effect | 133 | 247 | 0.188 | 0.247 | 0.76 (0.50, 1.15) | -- |
| Total effect | 1608 | 1699 | 0.236 | 0.264 | 0.90 (0.80, 1.01) | 0.88 (0.76, 1.01) |
| **Combined intervention**<br>(rfMDA + RAVC vs. RACD only) |  |  |  |  |  |  |
| Direct effect | 634 | 770 | 0.194 | 0.295 | 0.66 (0.54, 0.80) | -- |
| Spillover effect | 81 | 130 | 0.136 | 0.208 | 0.66 (0.55, 0.80) | -- |
| Total effect | 715 | 900 | 0.187 | 0.282 | 0.65 (0.34, 1.25) | -- |

**Table S10.** Cost-effectiveness analysis. Prevalent cases averted were estimated using hierarchical TMLE models for prevalence measured by qPCR. The number of prevalent cases averted equaled the produce of the difference in prevalence between arms among intervention recipients and non-recipients by the estimated population size within target areas vs. spillover zones. The incremental cost effectiveness ratio is the ratio of the difference in cost between arms by the difference in prevalent cases averted in both target area and spillover zones within 3 km of index cases for rfMDA + RAVC vs. RACD. Original estimates were reported in Ntuku et al., 2022 10.1136/bmjopen-2021-049050.

|  |  | N individuals |  | Prevalence |  | Prevalent cases |  | Total prevalent cases averted (95% CI) | Incremental cost-effectiveness ratio (95% CI) | % change from original estimate |
| --- | --- | --- | --- | --- | --- | --- | --- | --- | --- | --- |
|  | Intervention cost | Target area | Spillover zone | Target area | Spillover zone | Target area | Spillover zone |  |  |  |
| **Human intervention** |  |  |  |  |  |  |  |  |  |  |
| RACD | \$354,750 | 8,187 | 996 | 0.033 | 0.087 | 268 | 87 | (ref) | (ref) | |
| rfMDA | \$368,321 | 8,060 | 1,301 | 0.029 | 0.025 | 236 | 32 | 87 (77, 96) | \$156 (\$141, \$177) | -3% |
| **Mosquito intervention** |  |  |  |  |  |  |  |  |  |  |
| No RAVC | \$261,409 | 7,845 | 1,290 | 0.037 | 0.058 | 288 | 74 | (ref) | (ref) | |
| RAVC | \$461,661 | 8,426 | 980 | 0.026 | 0.051 | 217 | 50 | 95 (82, 108) | \$2,105 (\$1,859, \$2,430) | -21% |
| **Combined intervention** |  |  |  |  |  |  |  |  |  |  |
| RACD only | \$127,312 | 3,697 | 626 | 0.033 | 0.079 | 121 | 49 | (ref) | (ref) | |
| rfMDA+RAVC | \$234,223 | 3,878 | 635 | 0.017 | 0.017 | 66 | 11 | 94 (74, 113) | \$1,142 (\$944, \$1,446) | -37% |

## Supporting Information

### Study population

This study analyzed data from a cluster-randomized trial of focal malaria interventions conducted in Zambezi region of Namibia from January 1 to December 31, 2017 (NCT02610400) (1, 2). The region has seasonal malaria transmission that peaks between January and June.

*Plasmodium falciparum* is the dominant species, and annual *Pf* incidence was less than 15 per 1,000 from 2010-2015. In 2016, the incidence was 32.5 per 1,000 following an outbreak (3). In 2015, prevalence measured by loop-mediated isothermal amplification was 2.2% (4). In the study site, the Namibia Ministry of Health and Social Services routinely delivered case management and annual preseason household IRS with dichlorodiphenyltrichloroethane, with the exception of a small number of structures that were sprayed with deltamethrin. In addition, they offered reactive case detection (RACD) within 500 m of confirmed malaria cases, which included testing with rapid diagnostic tests and treatment with artemether-lumefrantrine and single-dose primaquine for those who tested positive.

### Cluster-randomized trial design

The trial included 56 clusters defined based on census enumeration areas that were within the catchment area of study health care facilities. Enumeration areas were eligible for inclusion in the trial if they 1) were located in the catchment areas of 11 health facilities, 2) had complete incidence data from 2012-13, and 3) had at least one incident case during the trial. Using a two-by-two factorial design, the trial randomized 56 clusters to four arms: 1) RACD only, 2) reactive focal mass drug administration (rfMDA) only, 3) reactive vector control (RAVC) + RACD, 4) RAVC + rfMDA. rfMDA included presumptive treatment with artemether-lumefrantrine to individuals in target areas (Extended Data Table 1). The trial used restricted randomization with the following criteria: mean annual incidence in 2013 and 2014, population size, population density, and mean distance from the household to a health-care facility. It was not practical to blind study participants or field staff to intervention assignment, but laboratory analyses and primary statistical analyses were blinded.

### Interventions

Field staff delivered interventions in response to passively detected malaria index cases that were confirmed by rapid diagnostic tests or microscopy if the case had resided in the study cluster at least one night in the prior 4 weeks. The trial delivered interventions in “target areas” within approximately 500 m of confirmed malaria cases detected through passive surveillance. In the RACD arms, individuals were eligible to receive rapid diagnostic tests, and individuals who tested positive were eligible for treatment with artemether-lumefrantrine and single-dose primaquine (Coartem, Novartis Pharmaceuticals, Kempton Park, South Africa; or Komefan 140, Mylan Laboratories, Sinnar, India). In the rfMDA arms, individuals were eligible for presumptive treatment with artemether-lumefrantrine. In the RAVC arms, households were eligible for IRS with pirimiphosmethyl (Actellic 300CS, Syngenta, Basel, Switzerland). In all arms, study teams aimed to deliver interventions within 500 m of a clinical malaria case and within 7 days to 5 weeks of the case report. RACD and rfMDA interventions were delivered to at least 25 people within target areas and RAVC was delivered to at least seven households within target areas.

Over 80% of eligible confirmed malaria cases received interventions, and over 85% of eligible intervention recipients were covered by interventions (2). Since compliance was high, for intervention recipients, we analyzed treatment as randomly assigned. Field staff did not offer repeat interventions in response to subsequent index cases within 5 weeks for rfMDA and RACD and within the same malaria season for RAVC. Field staff recorded the household geocoordinates of the index case and intervention recipients. Additional details about the interventions were previously published (1, 2).

### Procedures

Prior to randomization, field staff conducted a geographic census and recorded the latitude and longitude of all households in the study area. During the trial, trial staff extracted data on confirmed incident malaria cases and travel history from the rapid reporting system. At the end of malaria season between May and August 2017, the study team collected an endline cross-sectional survey to measure infection prevalence. Field staff collected dried blood spots on filter paper (Whatman 3 Corporation, Florham Park, NJ, USA) by finger prick from consenting individuals, and qPCR was performed targeting the acidic terminal sequence of the *var* gene.(5) Field staff also collected 250 ml of whole blood in BD Microtainer tubes with EDTA additive (Becton, Dickinson and Corporation, Franklin Lakes, NJ, USA) for serological analyses. Using human plasma, Luminex assays were performed to detect malaria antigens using previously described procedures (6, 7). Field staff recorded the geocoordinates of all sampled households.

### Informed consent

In the original trial, written informed consent was obtained from individual participants for rfMDA or RACD, and from heads of households (≥18 years of age) for RAVC. A parent or guardian was required to provide written informed consent for children younger than 18 years receiving rfMDA or RACD, and written assent for receiving these interventions was also obtained from children aged 12–17 years.

### Construction of analytic cohorts for incidence analysis

To construct cohorts, we matched index cases and intervention recipients to individuals recorded in the baseline census using household geocoordinates, age, and sex. We required that geocoordinates be < 100m apart to allow for small deviations in the location of geocoordinate recordings. We excluded 32 cohorts from the analysis for which it was not possible to merge intervention recipient geocoordinates with index data geocoordinates.

Because clusters were contiguous with no buffer zones between them, to capture potential dependencies across study clusters, we allowed cohorts to include individuals assigned to an adjacent cluster with a different treatment assignment from the triggering index case if it was within 1 km of an index case.

### Follow-up periods for analytic cohorts

We pre-specified cohort follow-up length based on the period in which we expected each intervention to reduce malaria among intervention recipients (direct effects) and non-recipients (spillover effects). Day 0 for each cohort was the date of index case detection. For comparisons of rfMDA and RACD interventions, the direct effect follow-up period was 0 to 35 days, the length of intrinsic incubation period for *Pf* malaria (8). This is the period of time in which we would expect the intervention to interrupt the parasite life cycle in treated, infected individuals, and in turn, prevent symptoms and/or infectiousness. The spillover effect follow-up period was 21 to 56 days; the 3-week lag period allowed for gametocyte clearance in the treated individual, sporozoite development in mosquitos, and development of detectable merozoites in humans.

For RAVC interventions, the direct effects follow-up period was 6 months since IRS can remain effective for an entire transmission season (9). The spillover effects follow-up period was from day 17 to 6 months. A mosquito bite could hypothetically be prevented on the day of intervention, so the earliest secondary case could occur after sporozoite development in mosquitos (minimum 10 days), and development of detectable merozoites in humans (minimum 7 days). We conducted a sensitivity analysis with alternative follow-up lengths (rfMDA and RACD direct effects: day 0-21; spillover effects: day 21-42; RAVC direct effects day 0-7; spillover effects day 17-90).

### Hierarchical TMLE

We compared incidence between arms using hierarchical targeted maximum likelihood estimation (TMLE) (10). We fit propensity score models at the cohort-level since interventions were delivered to cohorts. Within study clusters and cohorts, we expected individuals’ outcomes to be correlated due to interventions, social interactions, and local environmental factors. We fit two types of outcome models that accounted for statistical dependence in different ways (11). Cohort-level models allowed for statistical dependence between individuals in the same cohort without making any assumptions about the nature of the dependency.

Individual-level models assumed that cluster-level and individual-level covariates removed any dependence between outcomes of individuals in nearby geographic areas (11). We separately fit individual- and cohort-level models and then chose the outcome model with the smaller cross-validated mean squared error.

We fit outcome and propensity score models using an ensemble machine learning algorithm (the Superlearner) (12). For propensity score models, learners included generalized linear models, least absolute shrinkage and selection operator (LASSO) (13), and elastic net regression (14). For outcome models, we used the same learners as well as extreme gradient boosting (15). We performed 10-fold cross-validation using a loss function at either the individual- or cohort-level (11). Validation samples were constructed from randomly sampled individuals or cohorts. Because comparisons of rfMDA + RAVC vs. RACD had rare outcomes and a smaller sample size, we used 30-fold cross-validation.

### Adjusting standard errors for cohort overlap

We adjusted standard errors to account for potential correlation due to overlap between some cohorts using a model of cohort-level influence curves analogous to variance-covariance models used in cross-random effects models (16, 17). Specifically, we fit the model:

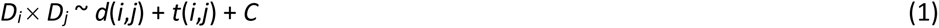

where *D_i_* × *D_j_* is the product of influence curves of cohorts *i* and *j*, *d*(*i*,*j*) is the distance between the location of the index case that triggered the intervention in each cohort, *t*(*i*,*j*) is the start date of the intervention in each cohort, and *C* is the cluster-level intervention assignment (18). Adjustment for intervention assignment accounted for correlation due to shared exposure to or receipt of the intervention. For cohorts with no overlap, we set *D_i_* × *D_j_*to zero. The regression was implemented with a simplified SuperLearner library including the generalized linear models and LASSO (13). We calculated the variance accounting for outcome dependence as follows:

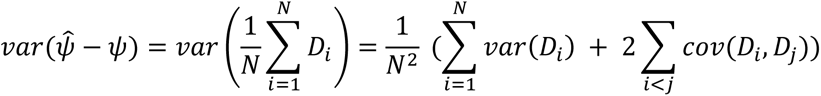

where *ψ̂* is the estimator, *ψ* is the estimand, and N is the number of cohorts.

In both incidence and prevalence analyses, we excluded any categorical covariates with less than 5% prevalence to avoid positivity violations. To minimize empirical positivity violations (19), we only fit models if the number of outcome events per variable was ≥10 and only fit adjusted models if the number of observations per strata was ≥30 (20).

### Deviations from pre-analysis plan

The analysis plan for this study was pre-specified at https://osf.io/s8ay4/. We note the following deviations from the plan:

1. We originally planned to conduct an individual participant data meta-analysis including data from three trials in Namibia, Eswatini, and Zambia. However, after reviewing the data for the Eswatini and Zambia trials, we determined that the geocoding of participants was not sufficient to allow for the planned spillover analyses. Thus, we proceeded with an analysis using data only from the Namibia trial.
2. In primary analyses using incidence data, we did not impose bounds on the mean outcome conditional on treatment and covariates because in initial models using bounds, estimates were very unstable.
3. In secondary analyses using prevalence data, we corrected standard errors at the cluster-level instead of at the household-level as specified in the pre-analysis plan. This better reflected the clustered sampling in the original trial.

## Notes

### Competing Interest Statement

The authors have declared no competing interest.

### Clinical Trial

NCT02610400

### Author Declarations

The trial protocol was approved by the Namibia Ministry of Health and Social Services (17/3/3) and the Institutional Review Boards at the University of California San Francisco (15-17422) and London School of Hygiene & Tropical Medicine (10411). The secondary analysis protocol was approved by the Stanford University Institutional Review Board (60708).

### Summary of Updates

We added Jennifer L Smith as co-author. She was accidentally omitted from the prior preprint.

